# The telemedical platform MyaLink for remote monitoring in myasthenia gravis: Rationale and protocol for a proof of concept study

**DOI:** 10.1101/2024.07.10.24310229

**Authors:** Maike Stein, Andreas Meisel, Maximilian Mönch, Pushpa Narayanaswami, Haoqi Sun, Meret Herdick, Lea Gerischer, Sophie Lehnerer

## Abstract

**Rationale:** Myasthenia gravis (MG) is a rare, chronic neurological disorder leading to fluctuating muscle weakness and potentially life-threatening crises. Patients often require life-long specialized treatment, but timely interventions are frequently hindered by the limited availability of specialists. Telemedical solutions at specialized centers enabling patient-physician interaction hold promise in bridging this gap, but are not yet available for MG. We developed ‘MyaLink,’ a remote monitoring platform tailored for MG, and outline the study design assessing the platform and clinical outcomes regarding telemedical intervention. Additionally, we present study results on care-related aspects in MG prior to telemedical intervention to identify challenges in the current care provision process.

**Design:** The platform comprises a patient app and a physician portal, enabling systematic symptom monitoring using data from patient-reported outcome measures (PROMs), coupled devices and a communication module. The randomized controlled study included 45 study participants (SP) over a 12-weeks period, including a group receiving standard care (15 MG patients) and a group with additional telemedical treatment (30 MG patients) including assessment of PROMs, wearable data collection and telemedical check-ups. Questions regarding care-related aspects were assessed at baseline visit.

**Results:** Many SP (N=33, 73.3%) communicate with the physician managing their MG via email. 73.3% (N=33) of SP identify areas for improvement in their MG care including symptom monitoring (N=23, 69.7%), specialist appointment availability (N=22, 66.7%), medication (N=22, 66.7%) and specialist accessibility (N=20, 60.6%). Additionally, 73.3% (N=33) reported that the effort required to manage their MG was high.

**Conclusion:** Our results emphasize the high demand of affected MG patients for continuous telemedicine services. MyaLink can provide such a service through personalized support based on the exchange of health data. Telemedicine solutions such as MyaLink promise to improve myasthenia care by providing accessible, patient-centred care that enables early detection of worsening symptoms and non-response to treatment.

**Trial registration:** The study was registered under DRKS00029907 on August 19, 2022.

## Rationale

Myasthenia gravis (MG) is a rare, chronic neurological disorder characterized by specific autoantibodies targeting the post-synaptic membrane of the neuromuscular junction. Symptoms present as fluctuating fatigability and weakness of ocular, bulbar, and skeletal muscles, potentially leading to life-threatening myasthenic crises [1]. Given its chronic nature, the majority of MG patients require long-term and often lifelong highly specialized care [2]. Yet much information about symptoms and individual disease conditions is lost between in-person appointments and across sector boundaries, and timely interventions are difficult due to limited access to specialists. At the same time there is a high individual counseling demand for MG patients on various topics. This is highlighted by a study analyzing >1700 patient inquiries sent by email to our specialized center and the German patient organization [3], as well as a comprehensive cross-platform validation of >2000 social media posts from MG patients [4]. This issue is further compounded by the recent approvals of targeted and costly intravenous and subcutaneous therapies (e.g., FcRn inhibitors, complement inhibitors) placing additional demands on both patients and healthcare professionals necessitating adaptations to healthcare structure and delivery. Telemedical solutions at specialized centers hold promise in bridging the gap in care accessibility by providing personalized support based on real-time health data. These solutions, which frequently use ‘patient-reported outcome measures’ (PROMs) for individual monitoring, are increasingly implemented in healthcare [5,6], also including various neurological disorders [7]. Yet their application in MG remains underexplored. Given that PROMs play an integral role in MG, both in clinical practice and research trials including large drug approval studies [8,9], monitoring disease activity and response to treatment, the field of MG is particularly well-suited for remote monitoring through digital technologies. To date, there are some studies investigating the utility of specific telemedical tools and scales for symptom monitoring in MG [10–16], also including machine-learning approaches for evaluation of remote symptom assessment [17–20]. However, no solution or study has yet facilitated or evaluated remote patient-physician interaction in MG. To address these needs, we have developed ‘MyaLink,’ a data-secure remote monitoring and patient-physician interaction platform specifically tailored to MG. Here, we introduce a new concept in the field of MG that uses longitudinal remote assessment as a tool to improve time-critical, needs-based care for MG patients, accessible through MyaLink. We outline the study design of the randomized controlled study evaluating the platform and present results from the study on care-related aspects in MG assessed during the baseline visit prior to telemedical intervention to identify challenges in the current clinical care provision process.

## Design

### Telemedicine platform and study design

#### Telemedicine platform

The telemedical platform utilized in the study (MyaLink) was developed in collaboration between the Charité Universitätsmedizin Berlin and the software partner Qurasoft GmbH. The patient organization Deutsche Myasthenie Gesellschaft e.V. (DMG) provided ongoing support and represented the patient perspective at regular meetings throughout all stages of development, ensuring a patient-centered approach. The platform comprises a patient application “app” (iOS, Android) and a web-based portal for physicians facilitating exchange of health data. Through the app, patients can monitor their symptoms using PROMs and assess vital parameters with coupled external devices (digital spirometer, activity tracker). A chat module enables patient-physician interaction for individual counseling. Patients can also store medical reports and documents in PDF format alongside a medication plan with a reminder function (**Figure 1**). The web-based portal enables physicians to review patient data in real time, providing an overview of the patient’s condition. Additionally, physicians can manage monitoring plans and adjust medications as necessary.

**Figure 1:**
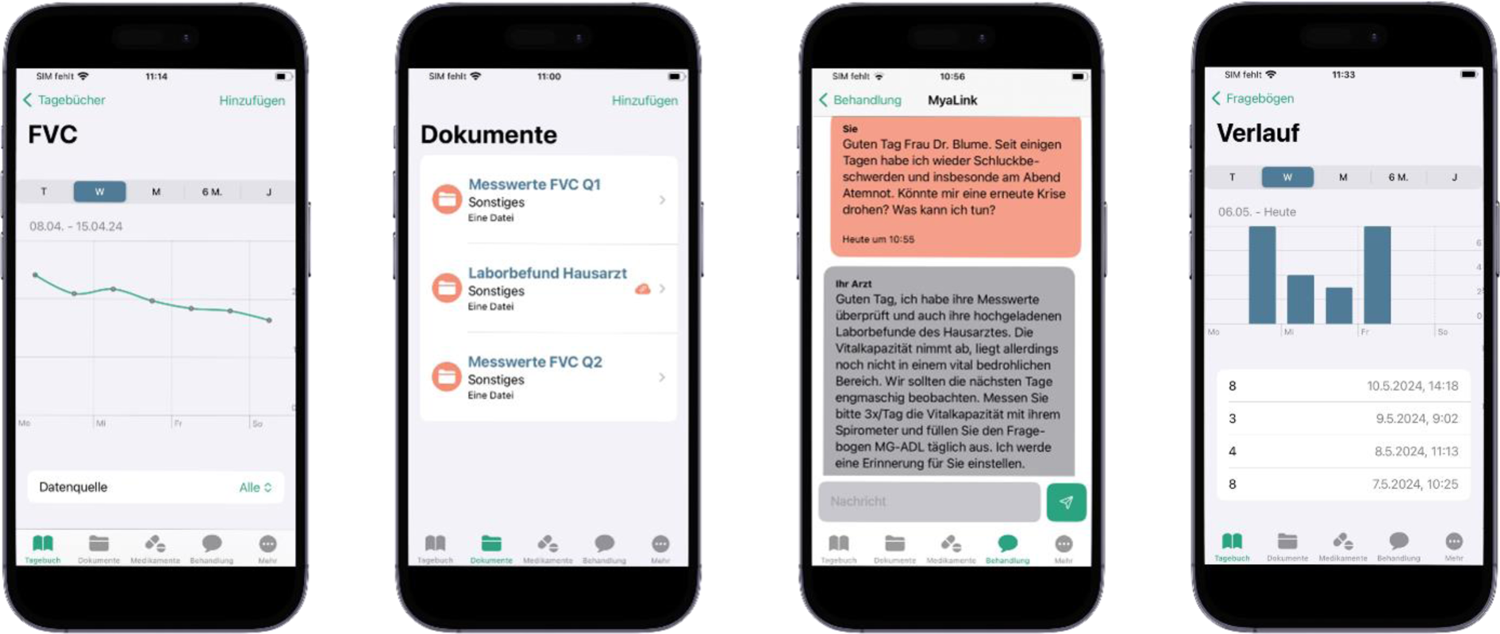
Modules of the MyaLink app (f.l.t.r.): (1) visualization of data entries for forced vital capacity (FVC) from spirometry (2) storage of documents (e.g., laboratory results, medical reports) (3) communication module and (4) assessment of PROMs (e.g., MG-ADL)

#### Data management

The platform is a certified CE marked (European Conformity certification indicating compliance with EU safety and performance standards) class-I Medical Device (MDD) and General Data Protection Regulation (GDPR) and Berlin Data Protection Act (BlnDSG) compliant. Data from the telemedicine platform is stored on certified data servers. Data transmission between patients and physicians is double encrypted using AES256. Data from visits at the study center and telemedical check-ups were documented electronically in REDCap® (Research Electronic Data Capture, Version 13.7.31).

#### Standard protocol approvals, registrations and patient consent

This study received approval by the ethics committee at Charité Universitätsmedizin Berlin (EA2/157/22). The study was conducted in accordance with the declaration of Helsinki. Study participants (SP) provided written informed consent prior to data collection/inclusion.

#### Eligibility criteria

To be eligible to participate in this study a SP had to meet the following criteria (recorded in the anamnesis):

Inclusion criteria:

- SP must have a diagnosis of MG (ICD-10: G70.0), for at least 6 months prior to enrollment
- SP must be undergoing treatment at the Charité Universitätsmedizin Berlin integrated myasthenia center (iMZ)
- SP must be able to communicate in German without an interpreter
- Age ≥ 18 years

Exclusion criteria:

- Organic brain diseases (ICD-10: F01-F09)
- Intellectual disability (ICD-10: F70-F79)
- Physical illnesses with limitations that make regular participation and data entry via a smartphone impossible
- Inability to conduct the study and understand the risks involved
- SP with somatic, psychiatric or neurological diseases or treatments that can impair cognitive functions or are not stable under drug treatment and could have an influence on the parameters to be examined
- SP who do not own a smartphone with an operating system of at least iOS 14 or Android 9

#### Recruitment, randomization, patient cohort

45 MG patients were recruited through our iMZ at Charité Universitätsmedizin Berlin. After enrollment and provision of written consent the SP were randomized in a 2:1 ratio into the intervention group (IG, N=30) or the control group (CG, N=15) using a digital randomization tool [21]. Randomization was stratified by sex and MGFA status classification at time of enrollment. SP in the intervention group could choose to undergo three additional Quantitative Myasthenia gravis Score (QMG) examinations [22] during the study period at the study center. Accordingly, they were assigned to either the IGQ-group (IG without additional QMG, N=12) or the IGQ+ group (IG with additional QMG, N=18). Due to the study design SP and physicians blinding was not possible.

#### Study design

The study consisted of a 12-weeks observation period between April and September 2023. All SP underwent two comprehensive visits at our study center including a clinical appointment at baseline and the end-of-study visit. During these visits, detailed data on MG-specific diagnostic results, clinical symptoms, comorbidities, medication, hospitalizations and care-related aspects in MG were collected. SP completed various PROMs, described in section ‘outcome measures’, and underwent an MG-specific physical examination (QMG). Additionally, the intervention group (IGQ+ and IGQ-) was equipped with a wearable (strapped on wrist activity tracker, Garmin Vivosmart 5) and a digital spirometer (MIR Spirobank Smart One). PROMs and reminders for spirometry measurements were assigned through the app at predefined intervals (weekly or monthly depending on the type of PROM/measurement). SP were instructed to wear the wearable during waking time, estimated as 14 hours per day, and optionally at night. Wearable and spirometry data was transmitted via bluetooth to the SP’s smartphone. At week 4 and week 8 physicians performed a telemedical check-up (TCU I and TCU II) including the review of monitoring data in the web-based portal. If necessary, SP were contacted through the chat by the physician (e.g., to change medication) based on the TCU results. SP could also initiate communication through the chat at any time, with physicians monitoring messages on weekdays and responding within 24 hours. SP in the IGQ+ group underwent three additional QMG assessments at the study center (week 3, 6 and 9). At the end of the study, the SP and healthcare professionals from the study team (N=5) completed usability questionnaires. A schematic overview of the study design is shown in Figure 2, and a detailed visit plan is presented in **Supplementary Table 1.**

**Figure 2:**
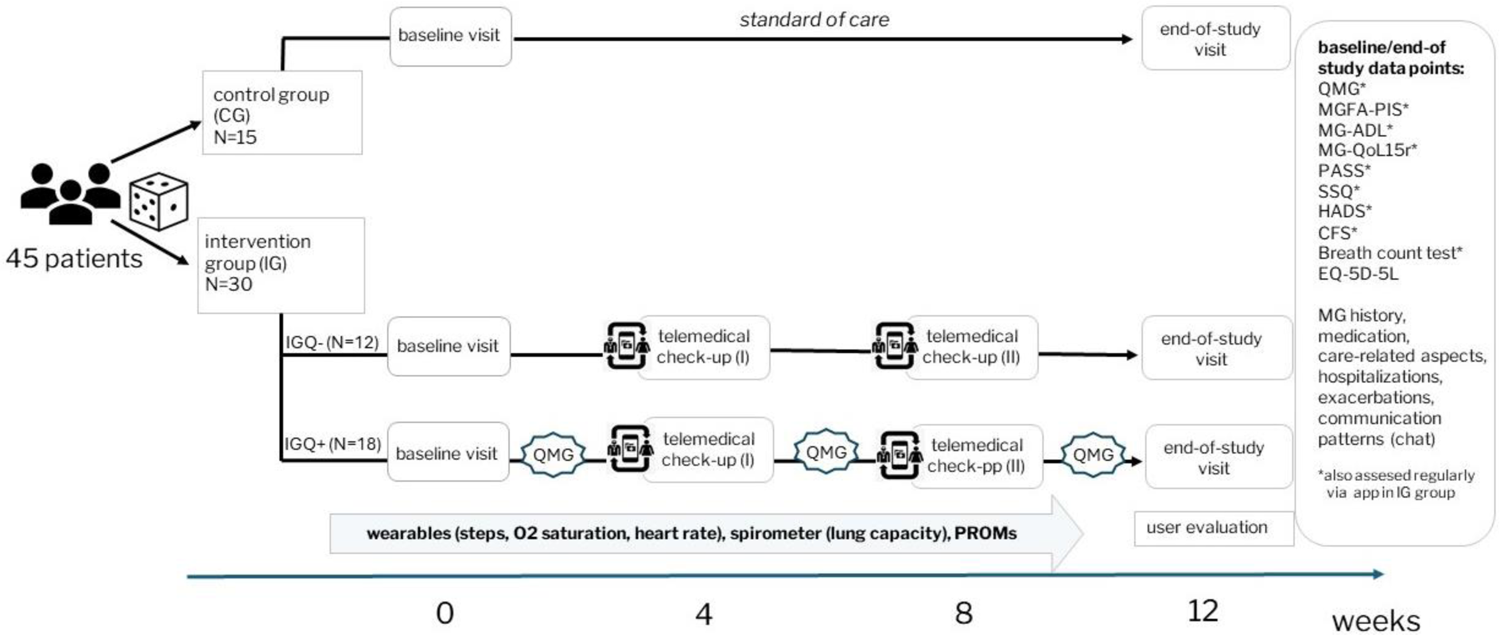
Schematic study design: 45 myasthenia gravis patients were randomized in a 2:1 ratio into either the intervention group (IG) or control group (CG). All study participants (SP) underwent a baseline and end-of-study visit at the study center. The intervention group, comprised of IGQ+ and IGQ-group, continuously monitored their symptoms throughout the study with external devices and completed patient-reported outcome measures (PROMs) assigned through the app. SP from the IG received telemedical check-ups conducted by study physicians in week 4 and week 8. Additionally, the IGQ+ group underwent three additional Quantitative Myasthenia gravis Score (QMG) examinations at the study center.

### Study objectives and outcome measures

The primary study objective was to assess the feasibility of digital assessment of PROMs and remote monitoring of vital parameters between treatment appointments in an iMZ.

Primary outcome measure:

- Adherence rates of collected PROMs defined as the ratio of completed questionnaires versus the number of assigned questionnaires through the app during the study period.

The secondary study objectives were to assess the usability of digital remote monitoring with the telemedicine platform from the perspective of SP and healthcare professionals, and to assess how this usability affects therapeutic alliance, empowerment, healthcare system utilization and clinical course.

Secondary outcome measures:

Assessment of usability:

- System Usability Scale (SUS) [23]
- Remote monitoring evaluation questionnaire (RMEQ): this questionnaire was adapted from Braun et al. [24] and specifically tailored to the MyaLink platform
- Dropout rate

Data from external devices (activity tracker, digital spirometer):

- O2 saturation (every minute), heart rate (every minute) and step count (hourly aggregated)
- Forced vital capacity (FVC)

Clinical endpoints (comparison of CG and IG group):

- QMG score [22]
- Single breath count test [25]
- MGFA status classification assessed at baseline visit [26]
- MGFA post-intervention status (MGFA-PIS) [26]
- Hospitalizations due to MG (including the type of ward and duration, therapies administered, potential triggers) and exacerbations (including the type of symptoms, potential triggers)
- Care-related aspects in MG

PROMs and questionnaires:

- Myasthenia gravis Activities of Daily Living (MG-ADL) [27]
- Myasthenia gravis Quality of Life, revised version (MG-QoL15r) [28]
- Chalder Fatigue Scale (CFS) [29,30]
- Hospital anxiety and depression scale (HADS) [31,32]
- Patient Acceptable Symptom State (PASS) [33]
- Single Simple Question (SSQ) [34]
- EQ-5D-5L [35]

A first analysis assessing feasibility and usability is available as a preprint [36]. Secondary endpoints are being analyzed. The following research questions will be evaluated in these analyses and presented in following manuscripts:

**1. Clinical endpoints analysis**: Does digital care with MyaLink reduce the disease severity of MG patients? Can MyaLink help to improve the quality of life of MG patients? The analysis will be exploratory using descriptive statistics comparing clinical endpoints between the control and intervention groups. Furthermore, effect sizes and confidence intervals will be reported. Results from this analysis will be used in follow-up studies for power calculations.
**2. Communication patterns and telemedical intervention analysis**: What are communication patterns and topics discussed between patients and physicians in the MyaLink chat? What insights can be gained from analyzing the telemedical check-ups regarding physician-initiated contacts and the resulting clinical implications? This analysis will include examining frequencies and topics of exchanged messages and the number of interaction days to explore communication behaviors. Furthermore, a qualitative evaluation of telemedical check-ups will be performed assessing the frequency and reason of physician-initiated contacts as well as the recommendations provided.
**3. Remote assessment of respiratory function in MG:** Is the single breath count single test a feasible tool for remote assessment of respiratory function in MG? Correlation analysis for the single breath count test and spirometry measurements will be performed.

### Care-related aspects in MG

#### Questions regarding care-related aspects in MG assessed at baseline

At the baseline visit, SP were asked questions regarding care-related aspects of their MG. These questions/statements encompassed areas such as the primary MG healthcare provider, their accessibility and communication channels utilized by SP to contact them. Additionally, SP were asked about areas they identified as requiring improvement in their MG care, along with their perceived level of influence over their MG.

#### Analysis strategy for care-related aspects in MG

Since this is a pilot study, a prior estimation of the effect sizes and power calculation was not possible. The statistical analyses were performed using R software (version 4.2.2) and R Studio software (version 2023.03.1 build 446) [37]. The assessment of care-related aspects in MG is summarized without group stratification by absolute and relative frequencies, not considering overlap in multiple response questions.

### Results of care-related aspects in MG

To evaluate the initial state of care-related aspects in MG, questions were directed to SP during the baseline visit prior to engaging in telemedical monitoring. 73.3% SP (N=33) stated their iMZ was their primary healthcare provider for MG and 20% SP (N=9) named their office-based neurologist. Preferences for communication with the physician managing their MG varied, with 73.3% (N=33) using email and 60.0% (N=27) favoring in-person consultations, whereas the combination of both ways was possible. The majority (N=33, 73.3%) of SP reported a wish for improvements in their MG care. Approximately two-thirds wanted enhancements in symptom monitoring (N=23, 69.7%), appointment availability (N=22, 66.7%), drug treatment (N=22, 66.7%) and specialist accessibility (N=20, 66.6%). Approximately half of those SP expressed a desire for improved management of comorbidities (N=17, 51.5%) and better coordination among healthcare professionals (N=16, 48.5%). 73.3% (N=33) of all SP considered the personal organizational effort required for the management of their MG to be high or very high. Views on personal influence over their MG varied, with 44.4% (N=20) stating they had no perceived influence over their MG. 55.5% (N=25) SP reported good MG specialist accessibility, 20.0% (N=9) SP indicated limitations and 24.4% (N=11) SP experienced poor accessibility (**Table 1**).

**Table 1:** Care-related aspects in MG. Questions were asked at baseline visit (N=45).

|  | n (%) |
| --- | --- |
| **Who is your primary healthcare provider for your MG?** |  |
| Primary care physician | 3 (6.7%) |
| Office-based neurologist | 9 (20.0%) |
| Specialist center (iMZ) | 33 (73.3%) |
| **How do you reach your primary healthcare provider? (multiple answers possible)** |  |
| Telephone | 12 (26.7%) |
| Email | 33 (73.3%) |
| In-person or during clinical appointments | 27 (60.0%) |
| Neurological ward | 1 (2.2%) |
| Instant messenger provider | 1 (2.2%) |
| **I believe there is a need for improvement in the management of my MG.** |  |
| Yes | 33 (73.3%) |
| No | 12 (26.7%) |
| **If yes:** |  |
| **I see a need for improvement in the following area(s): (multiple answers possible)** |  |
| Medication | 22 (66.7%) |
| Non-medication therapy (e.g., lifestyle recommendations) | 10 (30.3%) |
| Accessibility of a specialist (e.g., via email, telephone) | 20 (60.6%) |
| In-person appointment availability with a specialist | 22 (66.7%) |
| Coordination between different healthcare providers in regard to MG-related issues | 16 (48.5%) |
| Improved symptom monitoring | 23 (69.7%) |
| Treatment of comorbidities | 17 (51.5%) |
| Other | 1 (3.0%) |
| **I assess the overall time and organizational effort required to manage my MG as follows (e.g., appointments, travel to the specialist, etc.):** |  |
| Very high | 14 (31.1%) |
| Rather high | 19 (42.2%) |
| Not very high | 8 (17.8%) |
| Not high | 4 (8.9%) |
| **How do you evaluate the physical accessibility of your primary MG healthcare provider?** |  |
| Very good | 11 (24.4%) |
| Rather good | 14 (31.1%) |
| Not really good | 9 (20.0%) |
| Not good | 11 (24.4%) |
| **I feel helpless in my own sphere of influence over my MG.** |  |
| Strongly agree | 6 (13.3%) |
| I rather agree | 14 (31.1%) |
| I rather disagree | 14 (31.1%) |
| I do not agree | 11 (24.4%) |

## Discussion

We introduce MyaLink, a remote monitoring platform tailored to MG, facilitating time-critical needs-based care for MG patients based on remote monitoring and personalized support, and present the study design of a randomized controlled study assessing the feasibility of the platform and clinical outcomes in regard to telemedical intervention. Additionally, results from this study assessing care-related aspects in MG revealed a high demand for improvements in MG management as many SP reported considerable challenges due to non-standardized communication channels and limited access to specialists.

This aligns with a previous study revealing structural deficits in MG management with considerable geographical barriers resulting in significant travel distances for patients and caregivers to a specialized center [38]. MG patients experience a high burden of disease [39] and demonstrate pronounced individual counseling demand [3,4]. These findings and our results underline the high need for services that MyaLink can offer, particularly the good accessibility of specialists independent of time and location, one of the most critical unmet needs for people with rare diseases. This is in line with recommendations for telemedicine use in clinical care for MG [40,41] in accordance with national action plans for rare diseases, explicitly calling for integrating digital solutions into outpatient management [42,43].

By providing remote access to specialized expertise and continuous monitoring data, platforms such as MyaLink may be able to provide real-time insights into disease states enabling earlier detection of symptom deterioration and non-responses to treatment. In contrast to the current infrequent in-person visits such digital support systems allow timely interventions, improving treatment efficiency and deployment of available therapies, which could also help to avoid misuse of the healthcare system (e.g., unnecessary visits to emergency rooms). Moreover, telemedicine solutions like MyaLink, have the potential to foster a more patient-centered care, facilitating collaborative decision-making and supporting intersectoral cooperation. Further research is needed to evaluate their long-term implementation in clinical care and their potential for personalized treatment.

## DECLARATIONS

### Availability of data and materials

Data not provided in the article may be shared (anonymized) upon request from the corresponding author for purposes of replicating procedures and results.

## Author contributions

M.S. wrote the manuscript. M.S., S.L. and L.G. developed the study design, critically reviewed by A.M., M.S. and S.L. conducted the study. M.S., S.L. and L.G. interpreted the data. Statistical analyses were performed by M.M. A.M., M.M., U.G., P.N., H.S., M.H., L.G. and S.L. critically reviewed and edited the manuscript.

## Acknowledgements

We thank the NeuroScience Clinical Research Center at Charité Universitätsmedizin Berlin for their support in administration and throughout the formal processes essential for the successful execution of this study. The authors especially wish to thank Maja Olzewska, Josefina Kreusler, and Gabriela Jiron-Lozano for their support in establishing the REDCap® database. Furthermore, special acknowledgements are extended to Jens Brestrich and Dike Remstedt for their support in conducting the study. Additionally, the authors thank Qurasoft GmbH for providing the software for the study, namely Lale Zils from Qurasoft GmbH for the project assistance during the study and Sebastian Thunert, Lucas Keune and Ragnar Hirschbrich from the Qurasoft team for their technical support throughout the study. We are grateful for the participation of patients and for the many constructive discussions with DMG e.V. during the platform’s development and all study phases. Their feedback and suggestions were very valuable in improving the platform and ensuring a patient-centered perspective. We wish to thank all SP that participated in the study.

