## Supplemental Table 1 for "The telemedical platform MyaLink for remote monitoring in myasthenia gravis: Rationale and protocol for a proof of concept study"

### Supplementary material

Supplementary Table 1: Visit plan of randomized controlled study (N=45) with a study duration over 12 weeks.

| | week 0 | week 1 | week 2 | week 3 | week 3 | week 4 | week 5 | week 6 | week 6 | week 7 | week 8 | week 9 | week 9 | week 10 | week 11 | week 12 | |
|---|---|---|---|---|---|---|---|---|---|---|---|---|---|---|---|---|---|
| **visit/system** | base-line | app | app | app | study center | app/ physician portal | app | app | study center | app | app/ physician portal | app | study center | app | app | end-of-study | visit inde-pendent |
| **group** | CG, IG | IG | IG | IG | IGQ+ | IG | IG | IG | IGQ+ | IG | IG | IG | IGQ+ | IG | IG | CG, IG | CG, IG |
| inclusion, exclusion criteria | x | | | | | | | | | | | | | | | | |
| demographics | x | | | | | | | | | | | | | | | | |
| MGFA status | x | | | | | | | | | | | | | | | x | |
| MG history | x | | | | | | | | | | | | | | | | |
| medication | x | | | | | | | | | | | | | | | x | |
| hospitalizations | x | | | | | | | | | | | | | | | x | |
| care-related questions | x | | | | | | | | | | | | | | | x | |
| exacerbations | | | | | | | | | | | | | | | | x | |
| MGFA-PIS | | | | | | | | | | | | | | | | x | |
| wearables | | continuous assessment | | | | | | | | | | | | | | | |
| spirometry (FCV) | | x | x | x | x | x | x | x | x | x | x | x | x | x | x | x | x* |
| single breath count test | x | x | x | x | x | x | x | x | x | x | x | x | x | x | x | x | |
| QMG | x | | | | x | | | | x | | | | x | | | x | x* |
| **PROMs** | | | | | | | | | | | | | | | | | |
| MG-ADL | x | x | x | x | | x | x | x | | x | x | x | | x | x | x | |
| MG-QoL15r | x | x | x | x | | x | x | x | | x | x | x | | x | x | x | |
| SSQ | x | x | x | x | | x | x | x | | x | x | x | | x | x | x | |
| PASS | x | | | | | x | | | | | x | | | | | x | |
| CFS | x | | | | | x | | | | | x | | | | | | |
| HADS | x | | | | | x | | | | | x | | | | | | |
| EQ-5D-5L | x | | | | | | | | | | | | | | | x | |
| telemedical check up | | | | | | x | | | | | x | | | | | | |
| communication patterns | | | | | | x | | | | | x | | | | | x | |
| usability questionnaires | | | | | | | | | | | | | | | x | | |

x*: additional measurements or clinical data independent from study protocol was possible (e.g., when patient was hospitalized)
